# The importance of the timing of quarantine measures before symptom onset to prevent COVID-19 outbreaks - illustrated by Hong Kong’s intervention model

**DOI:** 10.1101/2020.05.03.20089482

**Authors:** Hsiang-Yu Yuan, Guiyuan Han, Hsiangkuo Yuan, Susanne Pfeiffer, Axiu Mao, Lindsey Wu, Dirk Pfeiffer

## Abstract

**Background:** The rapid expansion of the current COVID-19 outbreak has caused a global pandemic but how quarantine-based measures can prevent or suppress an outbreak without other more intrusive interventions has not yet been determined. Hong Kong had a massive influx of travellers from mainland China, where the outbreak began, during the early expansion period coinciding with the Lunar New Year festival; however, the spread of the virus has been relatively limited even without imposing severe control measures, such as a full city lockdown. Understanding how quarantine measures in Hong Kong were effective in limiting community spread can provide us with valuable insights into how to suppress an outbreak. However, challenges exist in evaluating the effects of quarantine on COVID-19 transmission dynamics in Hong Kong due to the fact that the effects of border control have to be also taken into account.

**Methods:** We have developed a two-layered susceptible-exposed-infectious-quarantined-recovered (SEIQR) meta-population model which can estimate the effects of quarantine on virus transmissibility after stratifying infections into imported and subsequent community infections, in a region closely connected to the outbreak’s source. We fitted the model to both imported and local confirmed case data with symptom onset from 18 January to 29 February 2020 in Hong Kong, together with daily transportation data and the transmission dynamics of COVID-19 from Wuhan and mainland China. After model fitting, epidemiological parameters and the timing of the start of quarantine for infected cases were estimated.

**Results:** The model estimated that the reproduction number of COVID-19 in Hong Kong was 0.76 (95% CI, 0.66 to 0.86), achieved through quarantining infected cases −0.57 days (95% CI, −4.21 − 3.88) relative to symptom onset, with an estimated incubation time of 5.43 days (95% CI, 1.30 − 9.47). However, if delaying the quarantine start by more than 1.43 days, the reproduction number would be greater than one, making community spread more likely. The model also determined the timing of the start of quarantine necessary in order to suppress an outbreak in the presence of population immunity.

**Conclusion:** The results suggest that the early quarantine for infected cases before symptom onset is a key factor to prevent COVID-19 outbreak.

## Introduction

The outbreak of coronavirus disease (COVID-19) caused by the SARS-CoV-2 virus was first identified in Wuhan, China, in December 2019, and was declared a global pandemic in March 2020 [1–3]. Regions in East Asia, such as Hong Kong, Taiwan, Korea and Japan, all faced an extremely high risk of community outbreaks due to the massive influx of travellers from mainland China to these countries during the Lunar New Year festival in January 2020 [4, 5]. Both Hong Kong and Taiwan have been thought to have the highest risk of COVID-19 outbreaks outside mainland China, however, compared to many other countries, the size of the outbreaks in these regions could still be considered limited up until now (April 2020) [6], suggesting that initial intervention policies have been successful [7, 8]. To understand how it was able to contain the COVID-19 outbreak could be contained in these regions is of great importance in order to limit the current global spread of the virus and to prevent future recurrent outbreaks.

Identifying COVID-19 intervention policies which are effective in containing community spread but without any potentially devastating socio-economic impacts is critically important [9]. Presently, non-pharmaceutical interventions, including suppression and mitigation strategies, have both been used to control the outbreak. Suppression strategies, such as transportation restrictions or city lockdowns [5, 10, 11], are often severe and, in most countries, it can be challenging to fully implement these repeatedly or long-term due to their profound socio-economic repercussions [12]. Mitigation strategies, for example, the current guideline for workers in critical industries proposed by the Centers for Disease Control and Prevention in the United States, focus on implementing precautionary measures, such as mask wearing or practicing social distancing at work, rather than quarantining exposed but asymptomatic cases [13]. Such a precautionary approach may only be sufficient to mitigate an outbreak but not stop it, given the evidence of the presence of pre-symptomatic transmission. [14–17].

Hong Kong’s intervention policies are largely focused on isolating COVID-19 cases and quarantining of close contacts of the cases together with medical surveillance of other contacts, regardless of whether they show symptoms or not, to reduce the risk of community transmissions [18]. The definition of a close contact of a COVID-19 case in Hong Kong refers to a person who has been in contact with the case two days before symptom onset following the national guidelines on COVID-19 prevention [19]. This approach requires that those persons (including critical workers) who had close contact with a case, should be quarantined (primarily quarantined at home, or at a designated facility), which is more restrictive than precautionary measures but with less severe socio-economic repercussions than a complete city lockdown. Due to the current high emergency response level in Hong Kong, all infected individuals under quarantine will eventually be identified and isolated in a hospital, and reported as confirmed cases, unless they show very mild or no symptoms, or test negative for COVID-19. How Hong Kong successfully prevented a community outbreak through quarantine measures while avoiding a city lockdown thus offers us a useful perspective on determining intervention strategies for other countries/regions.

Community acquired infections in Hong Kong were first detected immediately after infected travellers entered Hong Kong from mainland China at the beginning of the Lunar New Year festival, which lasts for 7 days from 24 January to January 30 in 2020, and transmitted the disease. During the festival, more than one million travellers (including Hong Kong residents) arrived in Hong Kong from mainland China in a single week [20, 21]. As in most other countries, border control and quarantine policies were implemented in order to stop the initial spread of COVID-19 from Wuhan to Hong Kong. Up until 4 February 2020, when most of the border crossings were closed, infected cases with a travel history from Wuhan or mainland China were reported almost daily and community acquired infections were subsequently detected as well. [22]. Due to the increasing number of the imported cases, starting on 28 January, travellers from mainland China meeting certain criteria were required to undergo 14 days self-quarantine. Compulsory quarantine was applied to all travellers from mainland China from 8 February. Surprisingly, up until the end of February, the total number of confirmed cases remained quite low without the exponential growth seen in many other countries, while public services such as public transportation were still running and businesses, shops and restaurants remained open.

The identification of the critical factors relating to quarantine measures and their effects on virus transmissibility in Hong Kong can provide valuable insight into how to implement quarantine measures to suppress a COVID-19 outbreak without profound severe socio-economic repercussions. However, there are several challenges related to estimating the effect of quarantine measures on COVID-19 transmission dynamics in Hong Kong. First, the information on which proportion of COVID-19 infected cases were quarantined and the timing of their quarantine start is generally not available, not only because these data may not be fully recorded but also because the true size of the infected population is not known. Therefore, these quantities have to be estimated using a modelling approach. However, due to the fact that many confirmed cases were imported cases from the epidemic regions rather than all cases being acquired locally, simple traditional transmission models, such as a susceptible-exposed-infectious-recovered (SEIR) model, were not sufficient to characterize the COVID-19 transmission dynamics properly in Hong Kong as these models treat all infections the same without considering their origins. To obtain the reproduction number, an indicator of the transmissibility of the virus, the average number of secondary infections of a given infected individual, needs to be estimated. If the proportion of imported cases is high, the changes in the total number of confirmed cases are not exclusively caused by those secondary infections acquired locally. As a result, the reproduction number cannot be estimated accurately using the simple SEIR model. Second, the number of imported cases can be greatly affected by the number of travellers entering Hong Kong and by border control measures. The number of travellers from mainland China to Hong Kong reduced substantially after most of the border crossings were closed on 4 February 2020 [22]. Hence, it is necessary to incorporate the effects of border control measures affecting transportation from mainland China to Hong Kong into the model.

In order to identify the critical components of the quarantine measures which were effective in controlling the spread of COVID-19 in Hong Kong, we aimed to develop a two-layered susceptible-exposed-infected-quarantined-recovered (SEIQR) meta-population model, embedded with passenger data from mainland China, that can stratify imported and local (community-acquired) cases to consider the effects of border control measures on the number of imported cases. This model can capture the transmission dynamics of both imported and local infections by estimating important epidemiological parameters including the reproduction number and the timing of the start of quarantine of an infected case relative to symptom onset. Furthermore, we will show how the model can be used to identify the timing of the start of quarantine which is necessary in order to suppress an outbreak in the presence of population immunity.

## Materials & Methods

### Data sources

We collected the date of symptom onset time for each newly infected case of COVID-19 from 18 January to 29 February 2020 in Hong Kong from the Centre for Health Protection, Government of the Hong Kong Special Administrative Region [23]. The daily number of newly infected cases of COVID-19 in Wuhan City and mainland China (excluding Wuhan) from 16 January to 29 February 2020 were collected from the National Health Commission of China [24]. Daily passenger data from mainland China during the corresponding period were obtained from the Hong Kong Immigration Department [21].

### Meta-population framework

A meta-population model framework was developed in order to consider the virus spreading from mainland China to Hong Kong. Assuming that the new emergence of COVID-19 causes an outbreak at location *i* (e.g. Wuhan or mainland China (excluding Wuhan)), the daily number of exposed imported cases before symptom onset at a different location *j* (e.g. Hong Kong) can be calculated using our meta-population framework with a mobility matrix *M*. The model first generated the transmission dynamics in mainland China and used the transmission dynamics together with transportation data to estimate the number of exposed imported cases in Hong Kong. We assumed that patients could only cross the border to Hong Kong before their symptom onset and then became exposed imported cases. We modified a simple SIR model to generate the number of newly infected cases that would reproduce the similar cumulative numbers of confirmed cases as observed in Wuhan and mainland China (excluding Wuhan) (Figure S1 and Figure S2). Please see the supplementary methods for the detailed descriptions.

The mobility matrix element *M_ji_* represents the mobility rate from source *i* to target *j*, which is calculated by dividing the daily number of passengers from an epidemic source region to a target region by the population size in the respective source region (Table S1). The proportion of passengers from Wuhan amongst all passengers from mainland China during the study period can be calculated using the International Air Transport Association (IATA) database [25]. We estimated that 2.92% of all passengers from mainland China to Hong Kong were from Wuhan. The detailed steps to calculate daily numbers of newly exposed imported cases are described in the *section Two-layered SEIQR model*.

### Two-layered SEIQR model

We developed a two-layered SEIQR model to include the dynamics of both imported and local cases in Hong Kong (Figure 1) and linked this model to the meta-population framework:

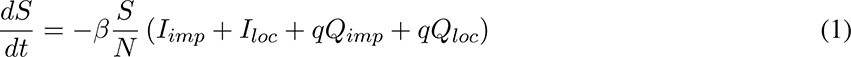

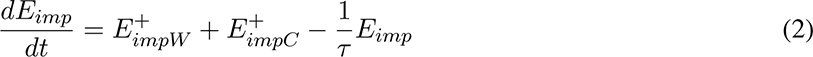

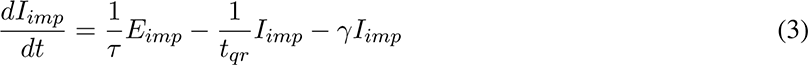

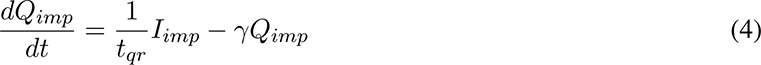

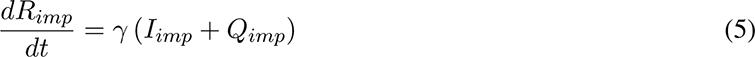

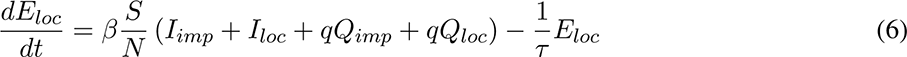

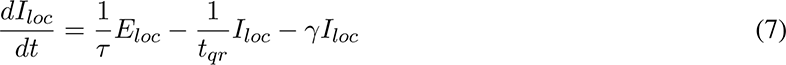

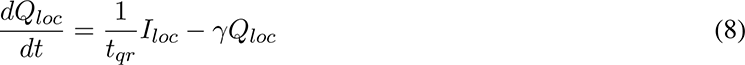

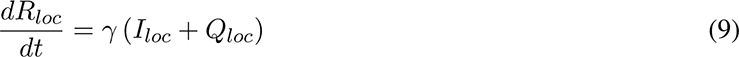

where *S*, *E*, *I*, *Q*, *R* represent the statuses of an individual: susceptible, exposed, infectious, quarantined, and recovered, subscripts *imp* and *loc* indicate imported and local cases, *β* is the transmission rate, *τ* is the latent period, *γ* is the recovery rate and *t_qr_* is the time interval between becoming infectious and the start of quarantine of an infected case and *q* is the ratio of the contact rates of quarantined to unquarantined patients. Note that, here the quarantined cases are infectious but are under quarantine. To avoid confusion, infectious cases only refer to infected individuals who are infectious but are not under quarantine. Also, we refer to all individuals, except susceptible, generated by our model as cases. Please see Table 1 and Table 2 for detailed variable and parameter definitions.

**Figure 1.**
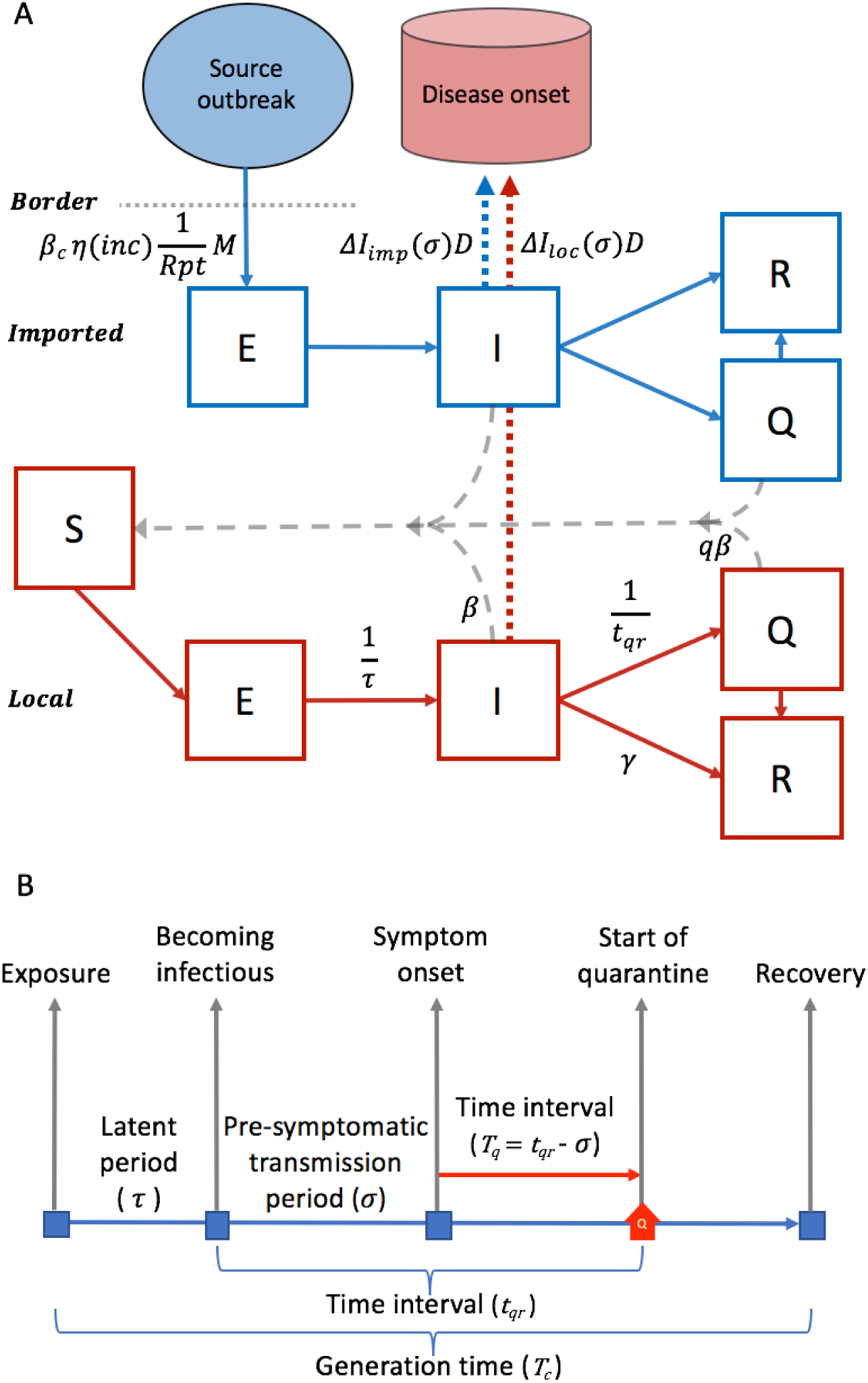
Model schema for transmission dynamics in Hong Kong with quarantine and border control measures. (A) Two-layered susceptible-exposed-infected-quarantined-recovered meta-population model. First layer (blue): Imported cases arrive with exposed (E) status before symptom onset to cross the border. Imported cases then become, sequentially, Infectious (I), Quarantined (Q) and Recovered (R). 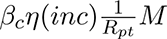 is the rate at which imported cases are produced from an epidemic source region (see Materials & Methods for the details). Second layer (red): Both imported and local infectious cases are able to infect susceptible individuals (S) and cause local (community) transmission. However, cases that are quarantined have a lower transmission rate than un-quarantined cases, defined by the ratio of the contact rates of quarantined to unquarantined patients *q*. *t_qr_* is the time interval between becoming infectious and the start of quarantine. Δ*I_imp_*(*σ*)*D* represents the number of newly detected imported cases, where *D* is the detection rate and *σ* is the pre-symptomatic transmission period, indicating the delay in time between becoming infectious and symptom onset. Similarly, Δ*I_loc_*(*σ*)*D* represents the number of newly detected local cases. For the definitions of epidemiological parameters, please refer to Materials & Methods. (B) Illustration of model parameters for an infected case. The house symbol indicates the start of quarantine. The red arrow refers to the time interval between symptom onset and quarantine start, which can be calculated as *T_q_* = *t_qr_* − *σ*. *T_q_* can be a negative value if the quarantine starts before symptom onset.

**Table 1:** Descriptions of variables in the meta-population model.

| **Variables** | **Descriptions** |
| --- | --- |
| $N$ | Population size in Hong Kong |
| $S$ | Susceptible individuals |
| $E_{imp}$ | Exposed imported cases |
| $I_{imp}$ | Infectious imported cases |
| $Q_{imp}$ | Quarantined imported cases |
| $R_{imp}$ | Recovered imported cases |
| $E_{loc}$ | Exposed local cases |
| $I_{loc}$ | Infectious local cases |
| $Q_{loc}$ | Quarantined local cases |
| $R_{loc}$ | Recovered local cases |
| $N_W$ | Population size in Wuhan |
| $S_W$ | Susceptible individuals in Wuhan |
| $I_W$ | Infectious cases in Wuhan |
| $R_W$ | Recovered cases in Wuhan |
| $N_C$ | Population size in mainland China (excluding Wuhan) |
| $S_C$ | Susceptible individuals in mainland China (excluding Wuhan) |
| $I_C$ | Infectious cases in mainland China (excluding Wuhan) |
| $R_C$ | Recovered cases in mainland China (excluding Wuhan) |

**Table 2:** Posterior estimates of reproduction numbers and other epidemiological parameters for COVID-19 transmission and control in Hong Kong. Mean values with 95% credible intervals are produced. The effective reproduction number *R_e_* is the expected number of secondary infections with quarantine measures. The basic reproduction number *R*_0_ is defined as the expected number of secondary infections without quarantine measures 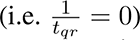 or population immunity. The incubation time *inc* refers to the sum of the latent period and the pre-symptomatic transmission period (*τ* + *σ*). The reproduction numbers *R_e_* and *R*_0_, and the time interval between symptom onset and quarantine start *T_q_* are derived from the estimated values of other parameters. Note that the transmission rate *β* is not displayed because the reproduction numbers along with other parameters represent the effect of the transmission rate.

| Parameters | Values | Descriptions |
| --- | --- | --- |
| $R_e$ | 0.76 (0.66 - 0.86) | Effective reproduction number |
| $R_0$ | 2.32 (1.19 - 4.42) | Basic reproduction number (without quarantine measures or population immunity) |
| $T_q$ | -0.57 (-4.21 - 3.88) | Time interval between symptom onset and quarantine start (unit: days) |
| $inc$ | 5.43 (1.30-9.47) | Incubation time (unit: days) |
| $\tau$ | 1.94 (0.34-4.17) | Latent period (unit: days) |
| $\sigma$ | 3.49 (0.48-5.80) | Pre-symptomatic transmission period (unit: days) |
| $Tc$ | 7.80 (6.81-8.79) | Generation time (unit: days) |
| $t_{qr}$ | 2.92 (0.65-7.81) | Time interval between becoming infectious and the start of quarantine (unit: days) |
| $q$ | 10.26% (5.32-17.8) | Ratio of the contact rates of quarantined to unquarantined patients |
| $R_{pt}$ | 22.46% (9.37-40.57) | Reporting ratio in mainland China (the percentage of infected cases that are reported) |

Our two-layered SEIQR meta-population model is embedded with daily passenger data from Wuhan and main-land China (excluding Wuhan). 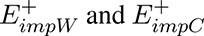 are the daily numbers of newly exposed imported cases from Wuhan (denoted by *W*) and mainland China (excluding Wuhan) (denoted by *C*) and are determined by the daily passenger numbers and incubation time:

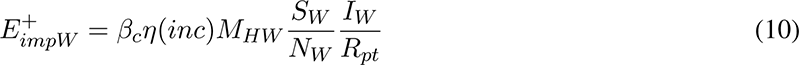

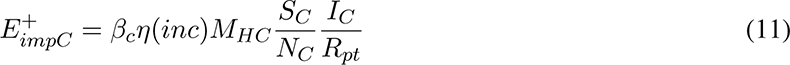

where *M_ji_* is the mobility rate from *i* to *j*, and subscripts *H*, *W* and *C* indicates Hong Kong, Wuhan and mainland China (excluding Wuhan), respectively. The terms 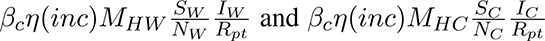 calculate the daily number of newly exposed imported cases from Wuhan and mainland China (excluding Wuhan), where *β_c_* is the transmission rate in mainland China, *η*(*inc*) is a function to calculate the number of infected cases prior to symptom onset (referred to the incubation time *inc*) [26] and *R_pt_* is the reporting ratio in mainland China, defined as the percentage of infected cases that are reported.

### Mapping model to symptom onset

The likelihood of observing symptom onset in both confirmed imported and confirmed local cases in Hong Kong was based on a Poisson distribution. The daily number of newly detected cases from the model was defined as the number of newly infected cases with symptom onset that had either been quarantined already or were quarantined after symptoms were detected, or were eventually isolated in hospital (without having been quarantined) after testing positive. Note that because the effects of quarantine and hospital isolation can be considered to be similar in terms of restrictions on movement to limit contacts in the outside world, patients in quarantine at home or at a designated facility, or in hospital isolation were all treated equally and given quarantine (*Q*) status to simplify the model mechanism. Thus, the definition of a newly detected infected cases was based on the assumption that all confirmed cases in Hong Kong were either quarantined or isolated. The cases that are recovered without having been quarantined or isolated are not detected. The daily number of newly detected imported cases with symptom onset was used as the expected value of the mean of the Poisson distribution and was derived as Δ*I_imp_*(*σ*)*D_imp_*, where 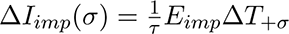, and Δ*T*_+_*_σ_* is a one day time step at a time delay between becoming infectious and symptom onset *σ*, referring to the pre-symptomatic transmission period. 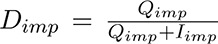 is the case detection rate of imported cases, which was defined as the proportion of the quarantined imported cases out of the quarantined imported and infectious imported cases (Figure 1A). This approach allowed the detection rate to be estimated. The same approach was used to determine the expected value of the mean of the Poisson distribution for the detected local cases as Δ*I_loc_*(*σ*)*D*, where 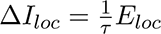.

### Effective reproduction number calculation

The effective reproduction number, *R_e_*, was calculated using the next-generation matrix approach after obtaining the posterior distributions of the model parameters [27]. Please see the supplementary methods for the detailed descriptions.

### Parameter estimation

The posterior distributions of the parameters of the SEIQR model for Hong Kong were obtained after fitting the model to the daily number of confirmed imported and local cases with symptom onset. The posterior distributions were estimated using a Markov Chain Monte Carlo (MCMC) algorithm with 1.2 × 10^6^ steps (Figure S3 and Figure S4) to guarantee an effective sample size (ESS) of greater than 200 for all parameters. The Gelman-Rubin convergence diagnostic was used and all scores were between 1 and 1.056, confirming that all parameter estimates successfully reached convergence.

Prior distributions for all parameters were set to uniform distributions, with the exception of the generation time *Tc* and the ratio of the contact rates of quarantined to unquarantined patients *q*. The generation time, defined as the average time interval between the infection time of an infected person and the infection time of the secondary infections, was implicitly assumed to be the sum of the infectious and latent periods of an infected case in the model [28, 29]. The prior of the generation time was normally distributed with a mean of 7.95 days, the average from two previous studies [30, 31], and standard deviation of 0.25. The prior of the ratio of the contact rates *q* also followed a normal distribution with a mean of 12% and a standard deviation of 0.05. A recent study by Kwok et al. estimated that each individual can contact an average of 12.5 persons during a day [32]. Assuming many home-quarantined individuals are likely to have contact with individuals in their own household (expected number of 1.5 persons on average), the mean ratio of the contact rates of quarantined to unquarantined patients in the prior distribution can thus be estimated as 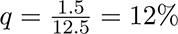.

## Results

### Dynamics of Imported and Community Cases

Our meta-population model was able to accurately reproduce the COVID-19 transmission dynamics of both imported and local infections in Hong Kong. The cumulative number of imported cases in Hong Kong increased rapidly after the first imported case was detected at the beginning of the Lunar New Year festival with the symptom onset day reported as 18 January 2020 (Figure 2A). Soon after, community-acquired infections started to occur. The risk of community spread was highlighted by the fact that the curve representing the number of local cases crossed above the curve of the imported cases. The model captured the cumulative numbers of local and imported cases, including the crossover of the curves (Figure 2B) and their transient dynamics (Figure 3). The predicted number of imported cases reached a peak on 26 January and soon reduced to near zero after 4 February due to border closures and a reduction in the number of outbreaks in mainland China (Figure 3A). These early imported cases immediately caused a wave of local infections. The daily number of newly detected local cases continued to increase and reached a peak after two weeks and community spread was suppressed soon afterwards (Figure 3B). During this period, prevention policies based on quarantine measures were implemented (Table S2).

**Figure 2.**
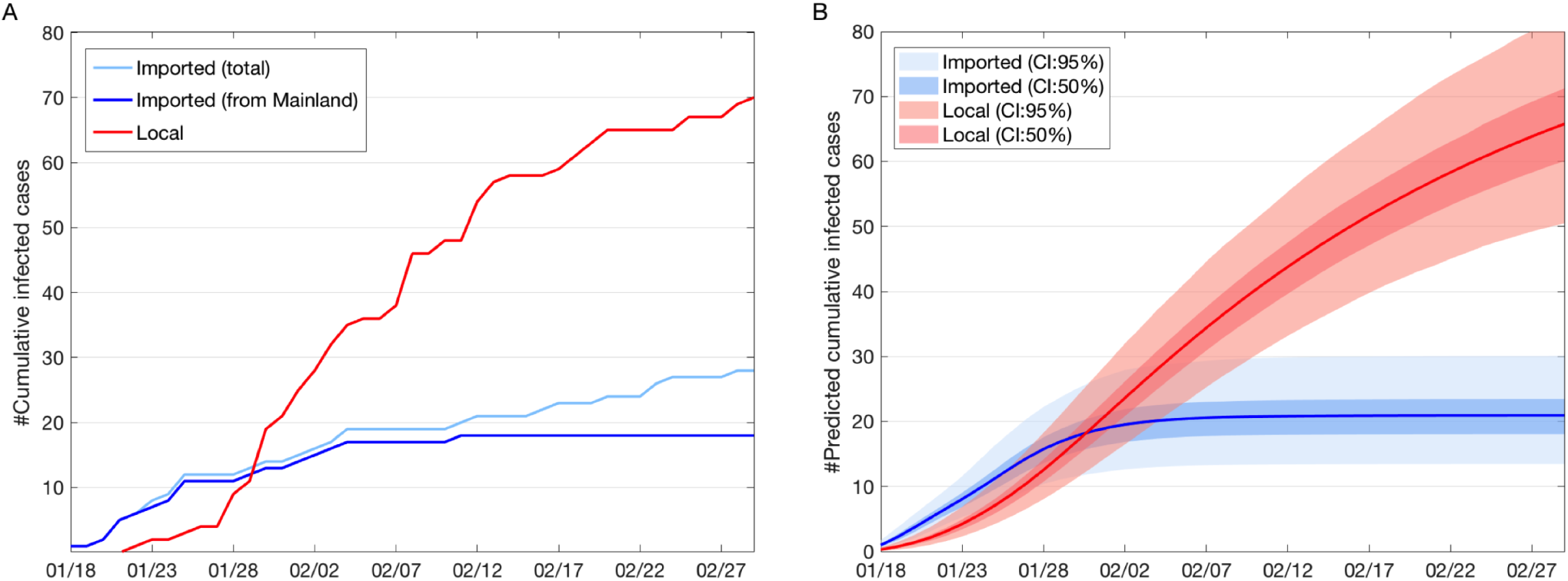
Cumulative number of confirmed COVID-19 cases by symptom onset date. (A) Observed cumulative number of infected cases. Light blue denotes the total cumulative number of imported cases. Dark blue denotes the cumulative number of imported cases only from mainland China, excluding some cases mainly from the Diamond Princess cruise. Red denotes the cumulative number of local (community-acquired) cases. (B) Model-predicted cumulative number of (detected) infected cases. Blue denotes the predicted cumulative number of detected imported cases from mainland China. Red denotes the predicted cumulative number of detected local cases. Solid lines indicate their mean values.

**Figure 3.**
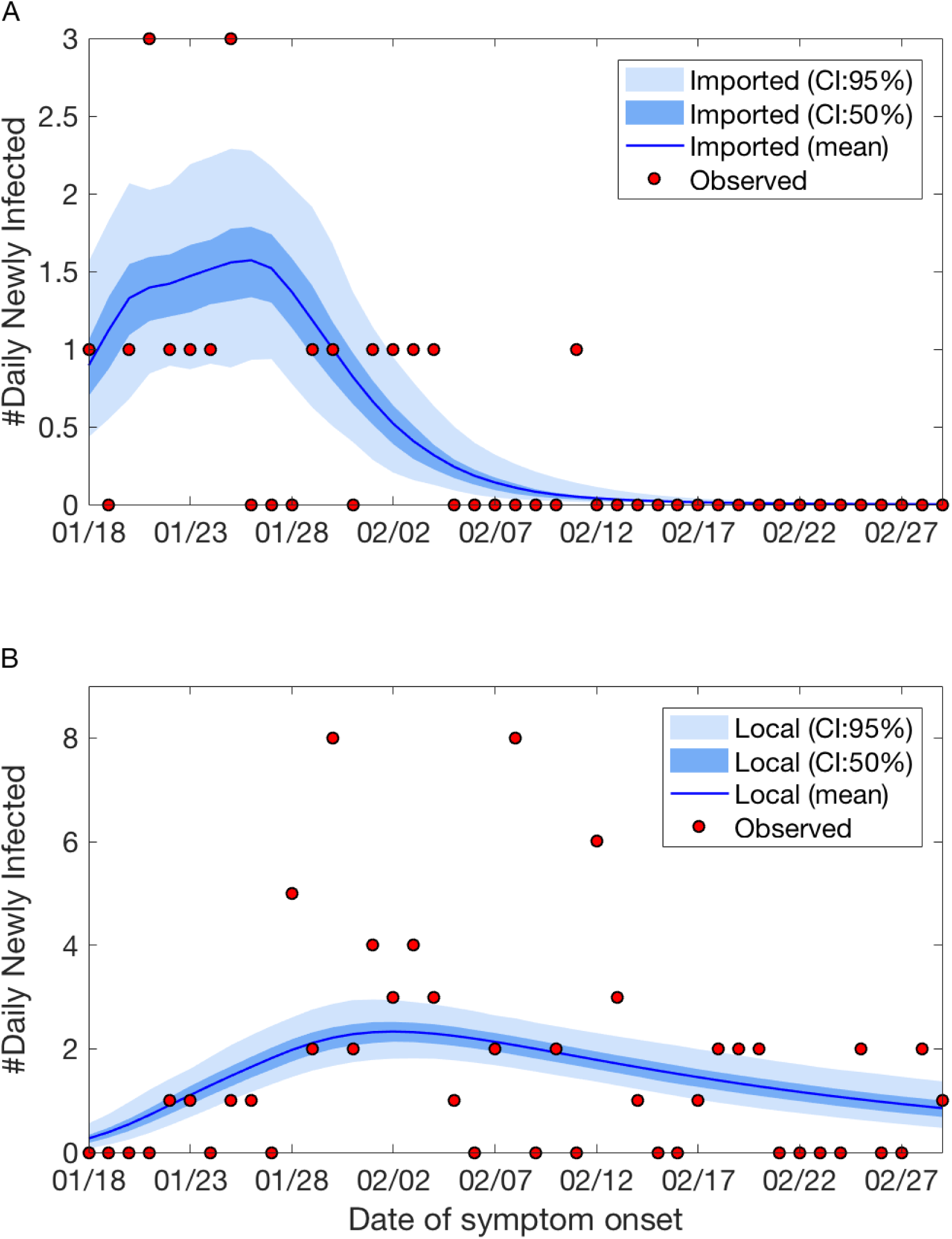
Observed and predicted numbers of detected imported and local cases. (A) Number of observed imported (red dots) and predicted detected imported cases. The solid blue line represents the mean model estimate, and the dark and light shading represent 50% and 95% credible intervals, respectively. The predicted detected cases are the daily newly detected cases with symptom onset that are quarantined or will eventually become quarantined, and have been confirmed positive. (B) Number of observed local (red dots) and predicted number of detected local cases. The solid blue line represents the mean model estimate, and the dark and light shading represent 50% and 95% credible intervals, respectively. The definition of the detected local cases is the same as in (A) but for local infections.

### Epidemiological Parameters

Epidemiological parameters of COVID-19 were estimated for Hong Kong from 18 January to 29 February. The effective reproduction number *R_e_* was 0.76 (95% CI, 0.66 - 0.86) (Table 2). The latent period was 1.94 days (95% CI, 0.34 - 4.17). The pre-symptomatic transmission period before symptom onset was 3.49 days (95% CI, 0.48 −5.80). The incubation time was calculated as 5.43 days (95% CI, 1.30 - 9.47), consistent with recent estimates for the mean or median incubation period of approximately 5 days [30, 33, 34]. The reporting ratio of infected cases in mainland China was estimated to be 22.46%(95%*CI*, 9.37 - 40.57). Sensitivity analysis was performed to evaluate the impact of generation time settings on the effective reproduction number. We tested two alternative model settings, with the generation time fixed at 7.5 [30] or 8.4 days [31]. These settings resulted in a mean *R_e_* of 0.76 or 0.77, respectively.

### Effects of Quarantine on Reproduction Number

The timing of the start of the quarantine of infected cases before their symptom onset is an important factor in stopping community spread. The time interval between symptom onset and quarantine start *T_q_* was obtained by subtracting the estimated time between becoming infectious and symptom onset (i.e. pre-symptomatic transmission period) from the estimated time between becoming infectious and the start of quarantine (Figure 1B). The time interval was used to represent the quarantine start time (relative to symptom onset) of an infected case. The estimated *T_q_* was - 0.57 days (95% CI, -4.21 - 3.88) (Table 2, Figure 4A), indicating that the low *R_e_* estimate was achieved through quarantining infected cases about half a day before their symptom onset. However, if *T_q_* was delayed by 1.43 days to 0.86 (95% CI, −2.78 - 5.31) days, i.e., less than one day after symptom onset, *R_e_* became greater than one, resulting in an increased risk of community spread. If no infected cases were quarantined, the reproduction number was calculated to be 2.32 (95% CI, 1.19 - 4.42), which was about 3-fold larger than the *R_e_* estimate in Hong Kong. This number was used as *R*_0_ in our study as *R*_0_, the basic reproduction number, is defined as the reproduction number in the absence of quarantine measures or population immunity. Furthermore, the ratio of the contact rates of quarantined to unquarantined patients *q* also affected the value of *R_e_*. The ratio *q* was estimated at 10.26%(95%*CI*, 5.32 – 17.80). If *q* changed to 25.27%(95%*CI*, 20.33 – 32.81), about two and a half fold increase, *R_e_* became greater than one resulting in potential community spread (Figure 4B). These results indicate that quarantining infected persons before symptom onset effectively can significantly reduce the risk of community spread.

**Figure 4.**
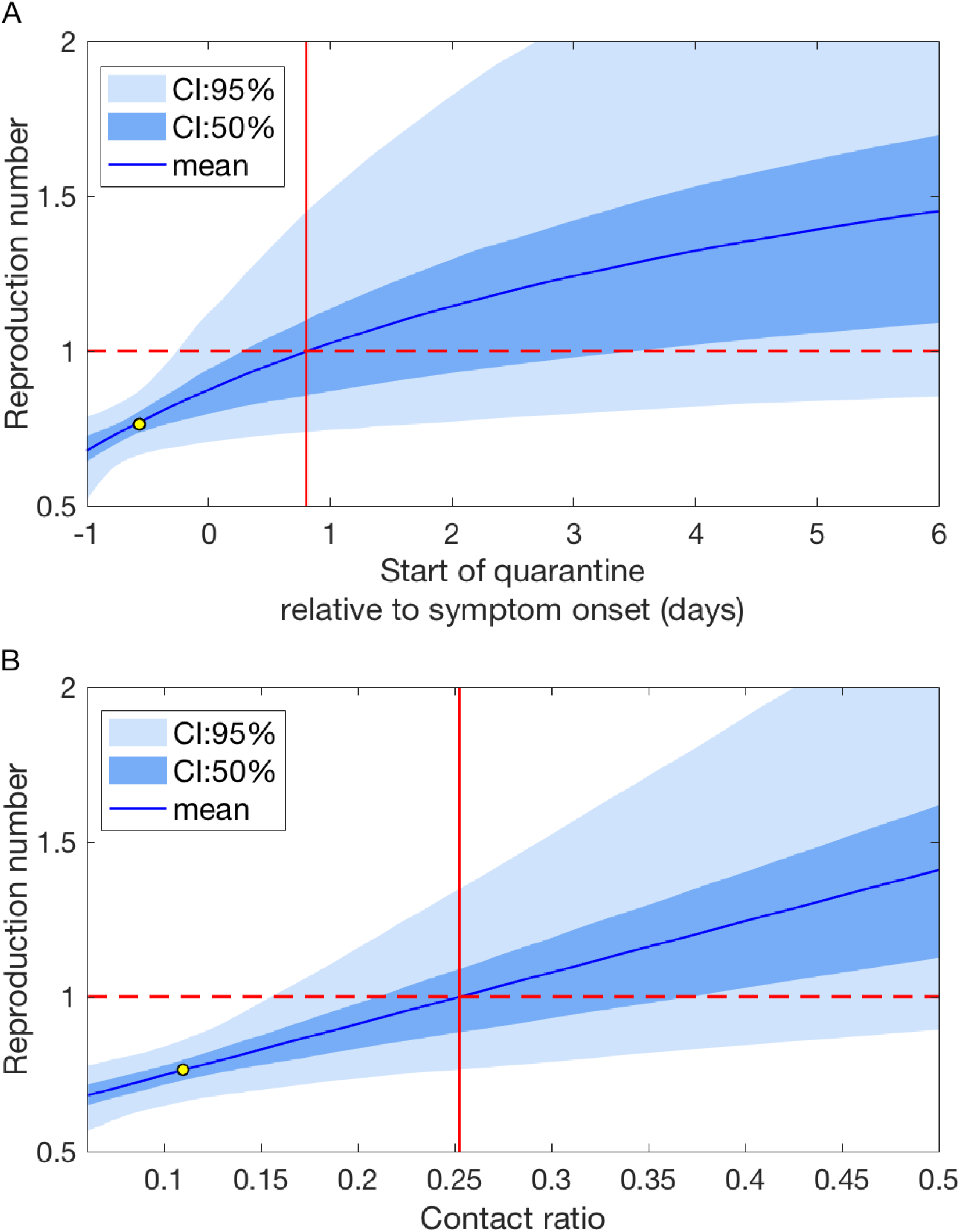
Effective reproduction numbers by different quarantine start times and contact ratios. (A) The mean and credible intervals of the effective reproduction number *R_e_* by quarantine start time *T_q_*. The vertical line refers to the average *T_q_* when *R_e_* is one. The dashed line indicates the level of *R_e_* = 1. The yellow dot represents the estimated *T_q_* and the corresponding value for *R_e_* in Hong Kong. (B) The mean and credible intervals of the effective reproduction number *R_e_* by the ratio of contact rates *q*. The yellow dot represents the estimated value of *q* and the corresponding value for *R_e_* in Hong Kong. All values were estimated from 10000 random samples from the posterior distributions.

We further studied the timing of the start of the quarantine which was critical for the suppression of the COVID-19 outbreaks in the presence of some level of population immunity under different social distancing scenarios. This critical timing was defined as the maximum difference in time between symptom onset and quarantine start that is able to reduce the reproduction number to one, which was used to represent the timing of the start of quarantine necessary in order to suppress an outbreak. We used *R*_0_, the basic reproduction number in a population without quarantine or population immunity to simulate various social distancing measures. Weak social distancing, as during the initial period in Wuhan, was assigned a high *R*_0_ of 3 ([35, 36]). Strong social distancing was represented by a low *R*_0_ of 2.32, adopted from our Hong Kong estimate when a large proportion of the population was vigilant about social distancing. Assuming a population immunity level of 30%, under weak social distancing the critical timing of the start of quarantine increased from 0.54 days before symptom onset to 1.72 days after symptom onset (Figure 5A), whereas under strong social distancing the average critical timing increased from 0.86 day to 5.85 days after symptom onset. If the quarantine started at the same time as the estimated timing in Hong Kong, the outbreak would be suppressed whether the social distancing is weak or strong (Figure 5B).

**Figure 5.**
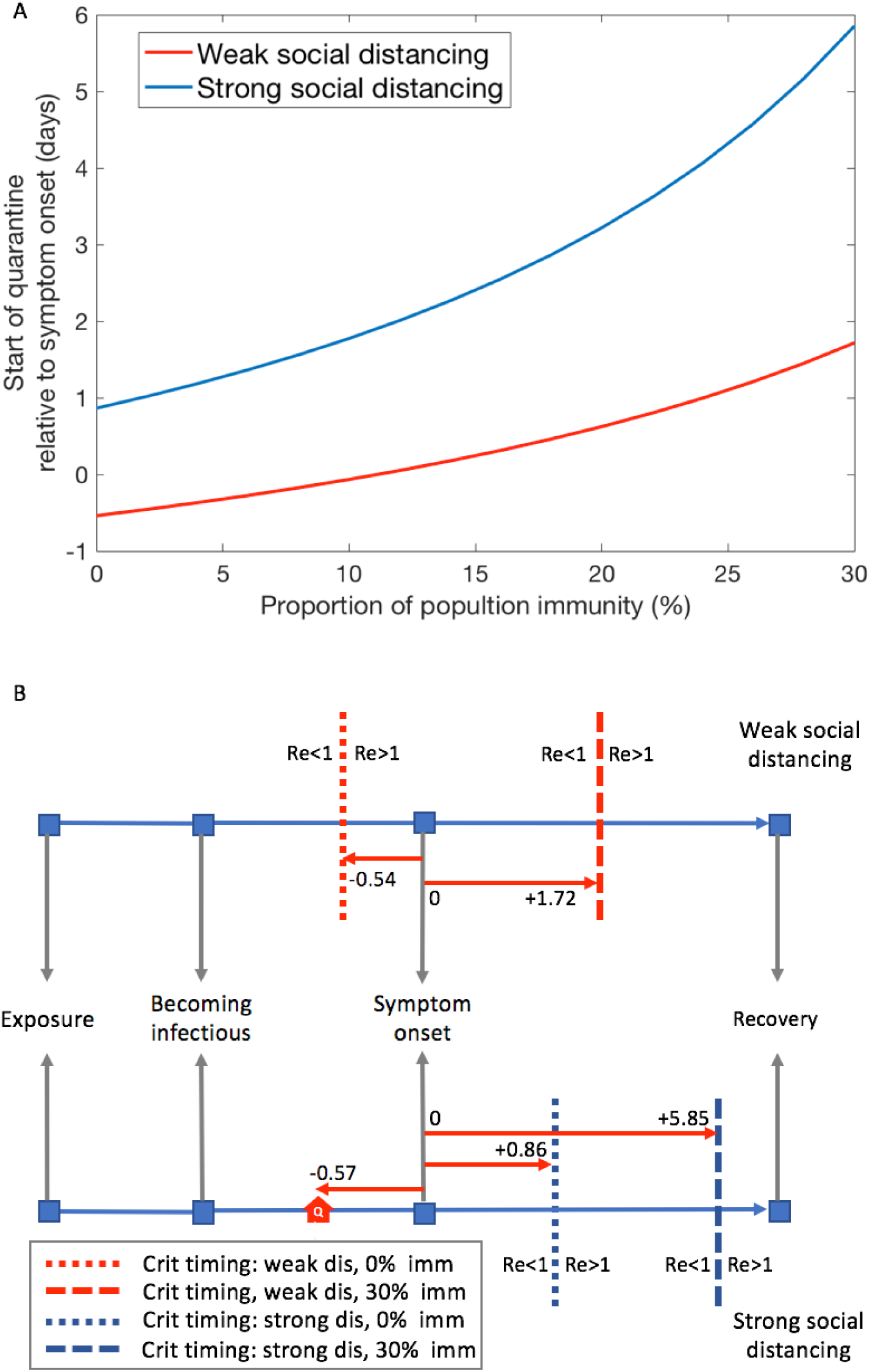
Time-sensitive quarantine measures under different social distancing scenarios. (A) Critical timing of the start of quarantine relative to symptom onset to suppress COVID-19 outbreak in the presence of herd immunity. The blue line indicates the timing of the start of quarantine to reduce the effective reproduction number *R_e_* to one with strong social distancing, corresponding to *R*_0_ = 2.32 (when quarantine effect is removed). The red line indicates the timing of the start of quarantine to reduce *R_e_* to one with weak social distancing, corresponding to *R*_0_ = 3 (when quarantine effect is removed). (B) Critical timings of the start of quarantine are illustrated within the timeline of an infected person from exposure to the virus until recovery. The upper panel shows the critical timing under weak social distancing. The red dotted line indicates the critical timing when population immunity is absent. The red dashed line indicates the critical timing at a level of 30% of population immunity. The lower panel displays the critical timing under strong social distancing. The house symbol indicates the estimated start of quarantine in Hong Kong. The definitions of the blue lines are the same as for the red lines in the upper panel.

### Effects of quarantine on Detection Rate

Early quarantine of infected cases allowed an increase in the detection rate of COVID-19 infections. The results showed that the detection rates reached stable values after three weeks from initially low values both for imported and local cases (Figure 6AB). Overall, the detection rates of the local cases were lower than those of the imported cases because the number of local infected cases increased faster than quarantined cases. Our model estimated that 71%(43 – 90) of local cases and 88% (58 - 99) of imported cases were detected at the end of the study period. However, a delay in starting quarantine of one day reduced the daily detection rate for local infections to 59%(38 – 74) (Figure 6B). Only 31%(20 – 40) of cases could be detected if quarantine was delayed by 6 days. The results showed that not only can early quarantine reduce the chance of transmission, but it also has the benefit of increasing the overall detection rate.

**Figure 6.**
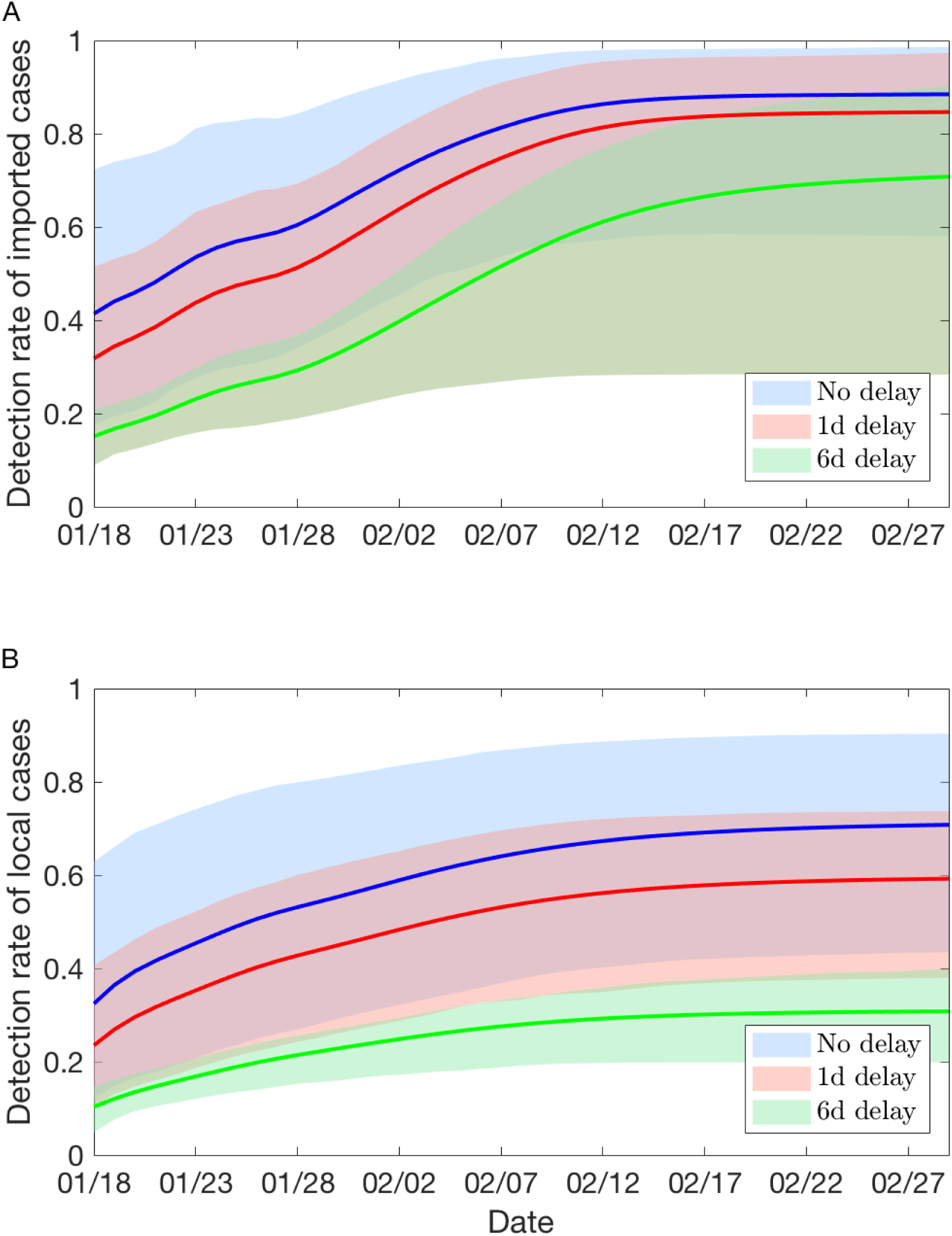
Detection rates of imported and local cases over time for different quarantine start times *T_q_*. (A) Detection rates of imported cases. Blue, the detection rate estimated using the estimate of the quarantine start time *T_q_*. Red, the detection rate estimated using the estimate of *T_q_* with one day delay. Green, the detection rate estimated using the estimate of *T_q_* with six days delay. The shaded areas represent the 95% credible intervals. (B) Detection rates of local cases. Same definition as for (A) are used for the estimates of local infections.

## Discussion

This is the first study to illustrate how Hong Kong can be a good model to learn how to prevent the community spread of COVID-19 through timely quarantine measures averting the need to adopt more intensive control efforts. By characterizing the disease transmission dynamics in Hong Kong during the early period of the global spread of the virus [10,37–39], our results demonstrated that an early quarantine of infected cases before symptom onset is a critical key factor in the suppression of COVID-19 community spread. After investigating Hong Kong’s intervention policies against community spread using our model, we propose time-sensitive quarantine measures based on the critical timing of the start of quarantine relative to symptom onset of infected cases to suppress the current or prevent future recurrent COVID-19 outbreaks, avoiding the significant socio-economic impact of more severe control measures as a consequence.

How to impose large-scale quarantine measures effectively to prevent or control the global expansions of COVID-19 without causing profound socio-economic repercussions remains largely unexplored. Until now, using strict intervention policies, such as transportation restriction or complete city lockdown, to suppress an outbreak of COVID-19 has been proposed and implemented in various locations [5,10,11]. These stringent policies can exert enormous socio-economic pressures with long-term consequences, not only now but also in future if recurrent outbreaks continue. One critical concern is that after the outbreak is suppressed, unless effective vaccines are available or a sufficient level of population immunity is achieved, an outbreak can reoccur after a short period of time and strict intervention policies will need to be reimplemented [12,40]. On the other hand, our model demonstrated that Hong Kong’s intervention policies can be used as a good example for achieving the suppression of community spread (defined by a reproduction number of less than one) or preventing any future recurrences through time-sensitive quarantine measures preempting the need for more restrictive control policies.

These time-sensitive quarantine measures represent an achievable way to suppress or prevent a COVID-19 outbreak. Ideally, quarantining every exposed person before they contact and potentially infect others can suppress the outbreak, but this strategy is not practically feasible since it either demands a lot of resources for tracing all possible contacts or may result in a de facto city lockdown situation as a large proportion of the population is quarantined. Another solution may be provided by implementation of a policy that aimed to time the start of quarantine for contacts of infected cases so that, the maximum difference in time between symptom onset and quanrantine start that is still able to reduce *R*_0_ to below one is achieved. This can be achieved by using contact tracing with modern technology for close contacts [7] and medical surveillance for persons who have potentially been exposed but are not classified as close contacts, as was done in Taiwan or Hong Kong. As the level of population immunity increases, the critical timing of the start of quarantine can be relaxed. Compliance with social distancing rules tends to weaken once the incidence of local cases is reduced, however, the proposed time-sensitive quarantine measures are adaptive to changes in population immunity level and social distancing compliance, which can provide public health authorities with an achievable and measurable target.

One of the major challenges to the implementation of any effective quarantine measure against COVID-19 is the possibility of pre-symptomatic transmission. Our model has found that an infected individual can be infectious without symptoms for about three days during the latent period. This result confirms the similar pre-symptomatic transmission periods found by several recent studies which were using contact tracing and enhanced investigation of clusters of confirmed cases [14–17] and gives reason to support an early quarantine for any persons who have been exposed but remain asymptomatic. Thus, it may be essential to expand the current definition of close contacts to include any contact with a confirmed case starting from 3-4 days before symptom onset [41].

To investigate quarantine measures in Hong Kong using a modelling approach is important in order to gain an understanding of the importance of the timing and the effects of quarantine under various scenarios. However, the difficulty lay in the characterization of disease transmission in order to obtain these effects where there is a mix of local and imported infections. We demonstrated that by using a two-layered disease transmission model that can stratify both the dynamics of imported and local infections, many epidemiological parameters, including the reproduction number, latent period and incubation time, can be correctly estimated after fitting the model to the symptom onset data of confirmed cases in Hong Kong. The resulting values are consistent with recent studies [30, 33, 34]. Another important reason for obtaining accurate estimates of many parameters in our model is that we assumed that only quarantined or isolated infected cases would be detected in Hong Kong. This assumption was made because, first, many infected cases were identified during quarantine and most infected cases were eventually isolated in hospital in Hong Kong; second, persons who had had contact with a case were requested to quarantine; third, mild or asymptomatic cases that were not quarantined were not likely to be detected. This assumption allows the model to fit symptom onset data of daily newly confirmed cases in Hong Kong well and to obtain the values for the timing of the start of quarantine and detection rate as well as other parameters.

Other interventions policies implemented in Hong Kong after the Lunar New Year holiday included the closure of schools and universities and the avoidance of unnecessary visits in elderly centres [22]. In addition, the proportion of healthy persons (including asymptomatic infected) wearing masks was high in Hong Kong during this period [42]. Although these interventions are not included specifically, our model results did not exclude the effects of these strategies. An estimated *R*_0_ of 2.32, which is lower than the value of *R*_0_ estimated during the initial periods of COVID-19 in mainland China [35, 36], can reflect the overall effects of these intervention policies. Since these effects aimed at reducing either transmission rates or contact rates, a lower *R*_0_ will be calculated without affecting other parameters. Our results suggest that adopting the quarantine measures implemented in Hong Kong can successfully limit community spread in the presence of other interventions.

The study illustrates the importance of quarantining exposed individuals early, about half a day before symptom onset to prevent or suppress a current COVID-19 outbreak without profound socio-economic impacts. Comparing the current timings of the start of quarantine of COVID-19 infected cases with the critical timing the model produced allows an assessment of the effectiveness of the quarantine measures. How to shorten the time from an individual becoming infectious to the the start of quarantine, so it leads to a substantial reduction of the reproduction number becomes the next challenge in working towards prevention and control of the current and future COVID-19 outbreaks.

## Data Availability

All data is available upon request.

## Acknowledgments

We thank Prof. Steven Riley from MRC Centre for Global Infectious Disease Analysis at Imperial College London to give valuable comments. We also thank Profs. Mengsu (Michael) Yang, Kai Liu, Xin Wang from City University of Hong Kong, Prof. Kin On Kwok from Chinese University of Hong Kong, Prof. Dong-Ping Wang from Stony Brook University, Prof. Jen-Tsan Ashley Chi from Duke University, Dr. Joey Leung from Georgia Institute of Technology, and all the anonymous readers who have provided invaluable comments. The authors also acknowledge the support from the grants funded by City University of Hong Kong [#7200573 and #9610416].

## Author Contributions

H-YY designed the study. AM and GH participated in the data collection. H-YY and AM analysed and interpreted the data. H-YY wrote the paper. Everyone reviewed, revised and edited the manuscript.

## Supplementary Methods

### Dynamics in source regions

To obtain the number of imported cases, the model has to generate the transmission dynamics in the source regions to seed the target region (Hong Kong). We modified an SIR model to construct number of newly infected cases that were close to the observed confirmed numbers in Wuhan and Mainland China (outside Wuhan):

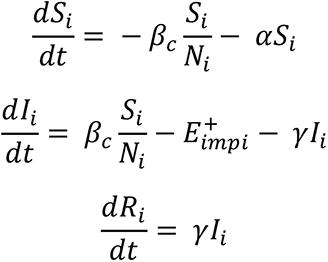

where S, I, R represent the statuses of an individual: susceptible, infectious, and recovered, subscript *i* indicates the source regions, *α* is a parameter to represent the effect of the reduction in social contacts after the Wuhan lockdown on 23 January 2020 and the closure of the border crossings on 4 February 2020 [1]. As a result, a reduction in the susceptible population happens by time. Assuming the generation time is 8.4 days, we have the recovery rate 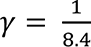. Using *R*_0_ = 2.92, we can obtain *β_c_* = *R*_0_*γ* = 0.3476 in Mainland China.

### Effective reproductive number calculation

The effective reproductive number *R_e_*, was calculated using the next-generation matrix approach after obtaining the posterior distributions of model parameters, following the same notation as in the study by Diekmann et al [2]. We obtained the transmission matrix *T* and the transition matrix *S*. The elements in *T* represent the average number of newly infected cases in the exposed group (E) caused by a single infected individual from the infectious (I) or quarantined group (Q), which can be calculated as 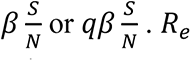 can be calculated as the first eigenvector of −(*TS*^−1^) with the following formulas:

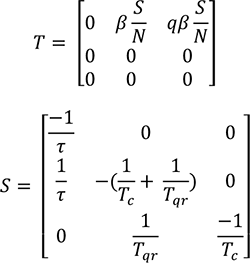

### Supplementary Figures and Tables

**Figure S1.**
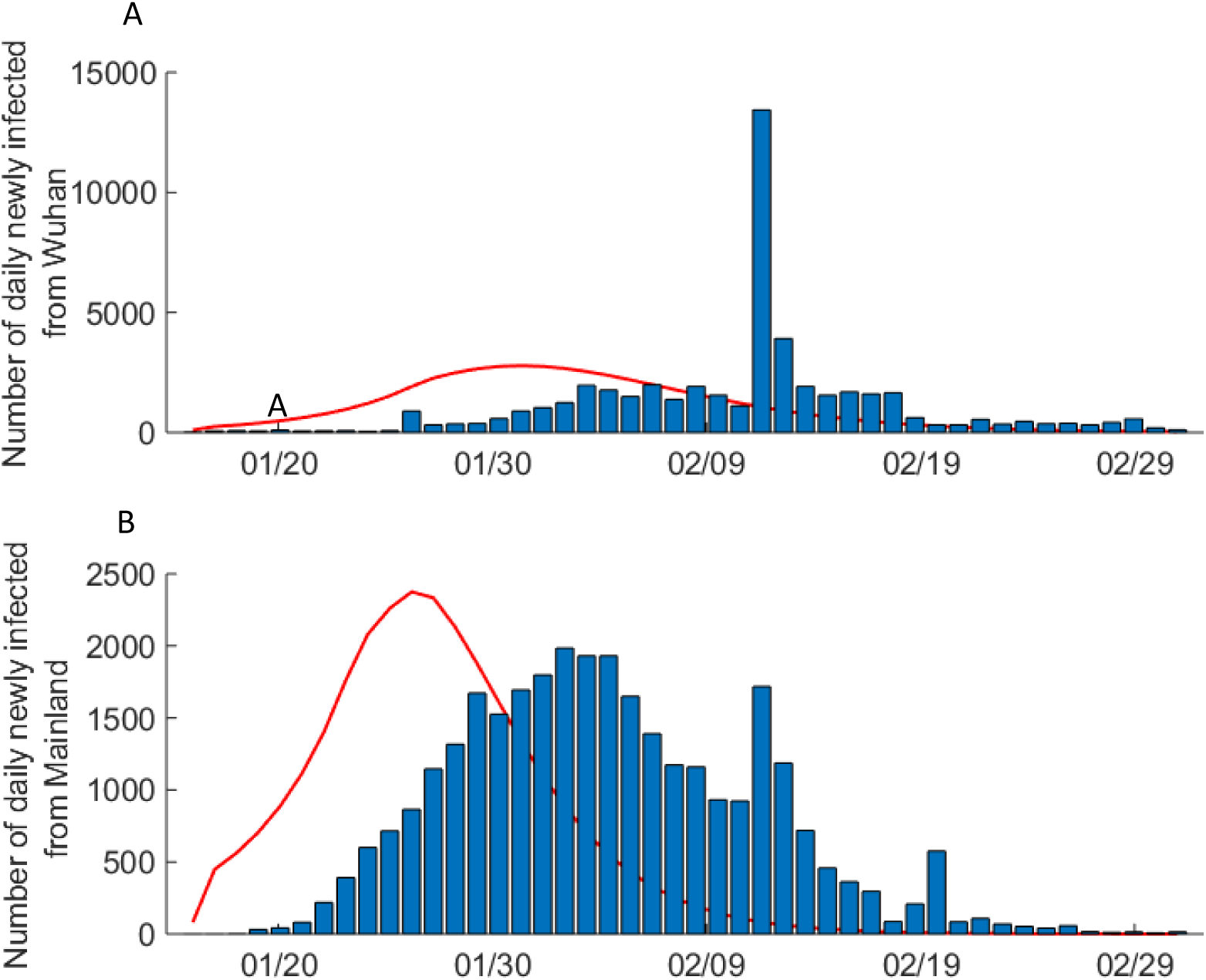
Daily number of newly infected COVID-19 cases in China. (A) Daily number of newly infected COVID-19 cases in Wuhan, China. The red line represents the reconstructed curve of the newly infected COVID-19 cases adjusted by a 10 day reporting delay after fitting the actual data from Wuhan, China. The x-axis denotes the case reporting date after Jan 16, 2020. (B) Daily number of newly infected COVID-19 cases in Mainland China (excluding Wuhan). The red line represents the reconstructed curve of the newly infected COVID-19 cases adjusted by a 10 day reporting delay after fitting the actual data from mainland China (excluding Wuhan).

**Figure S2.**
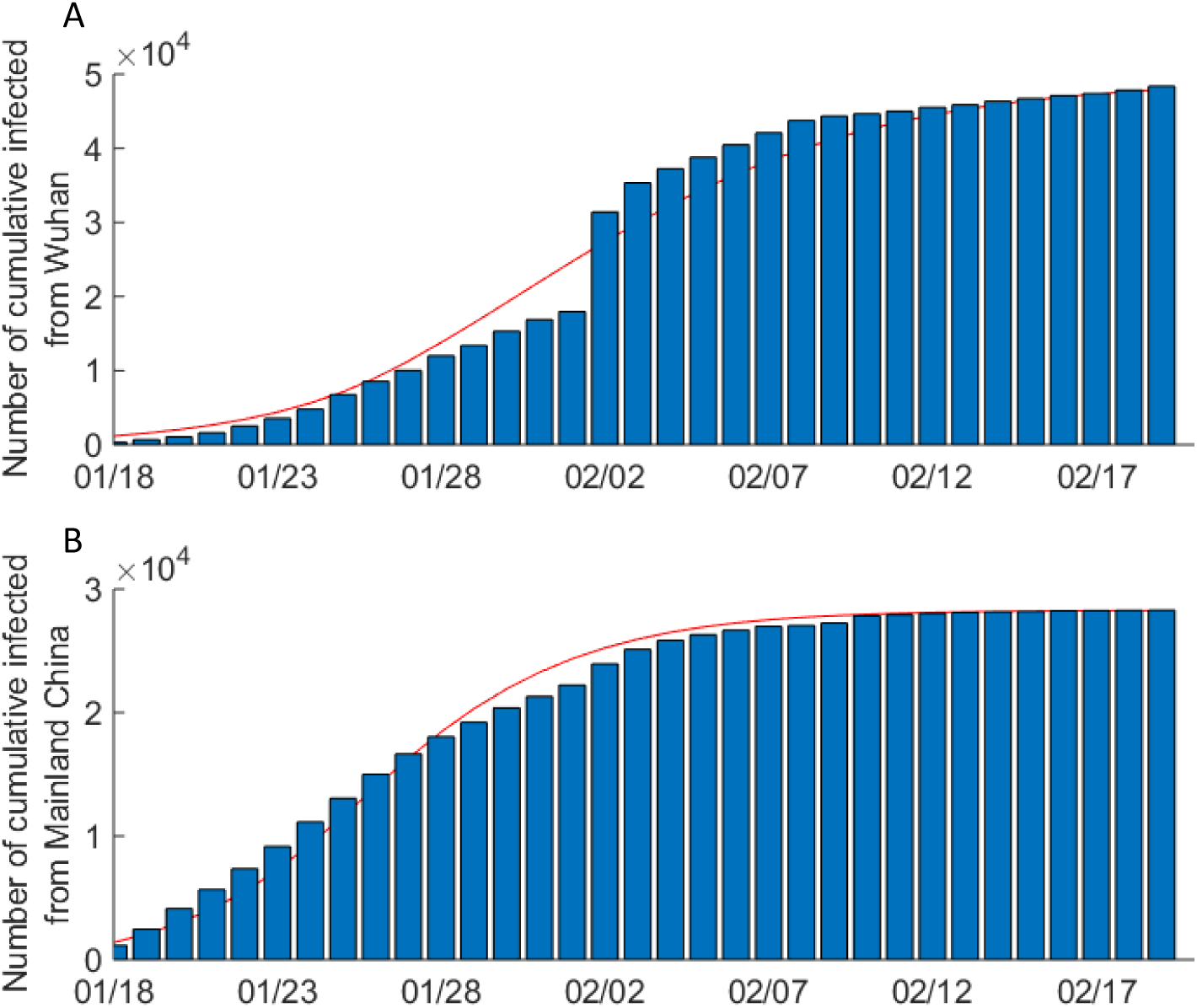
Cumulative number of confirmed COVID-19 cases in China. (A) Cumulative number of confirmed COVID-19 cases in Wuhan, China. The red line represents the reconstructed curve after fitting the actual data from Wuhan, China. The x-axis denotes the number of days from Jan 18 to Feb 29, 2020. (B) Cumulative number of confirmed COVID-19 cases in Mainland China (excluding Wuhan). The red line represents the reconstructed curve after fitting the actual data from mainland China (excluding Wuhan).

**Figure S3.**
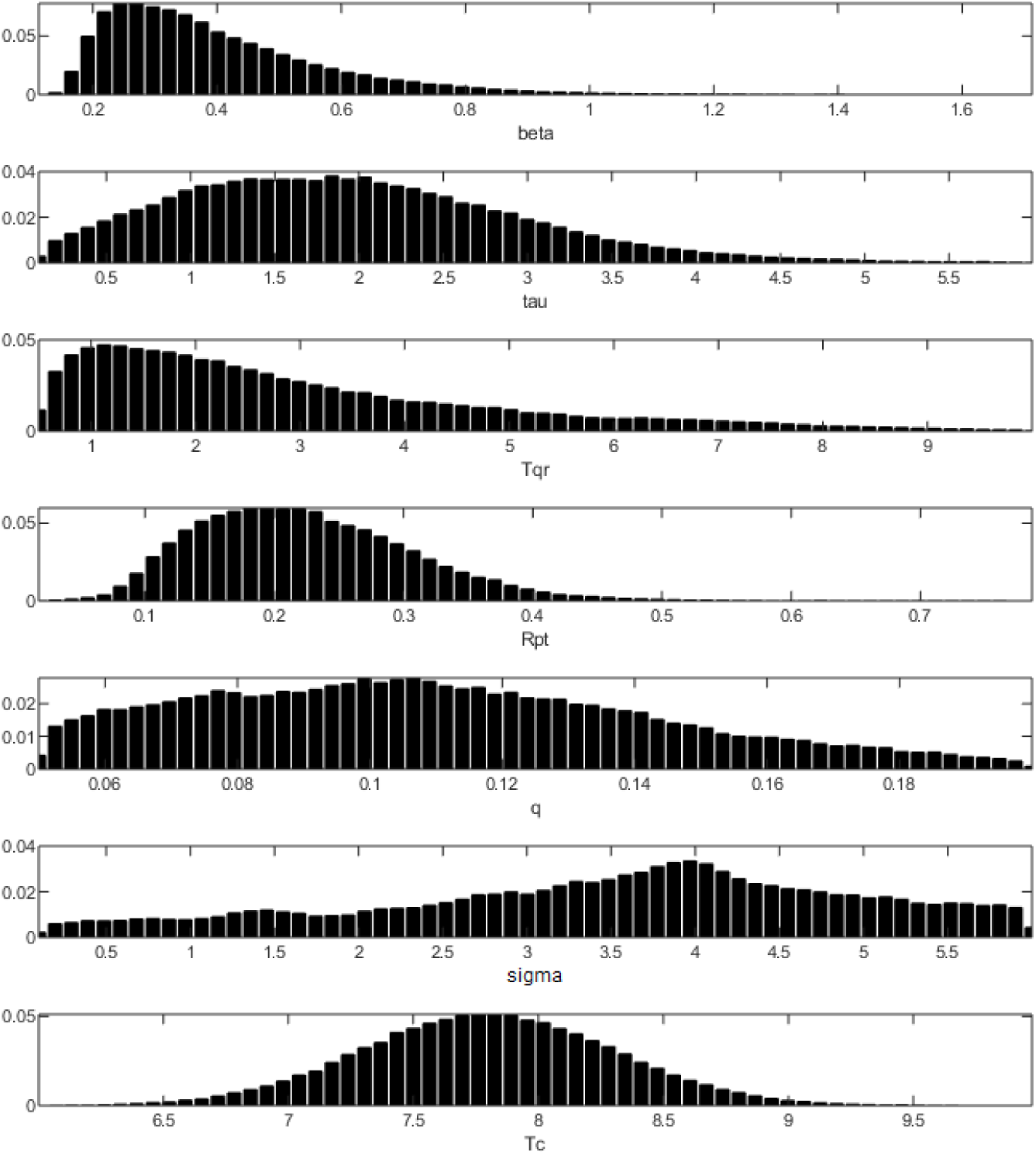
Posterior distributions of model parameters.

**Figure S4.**
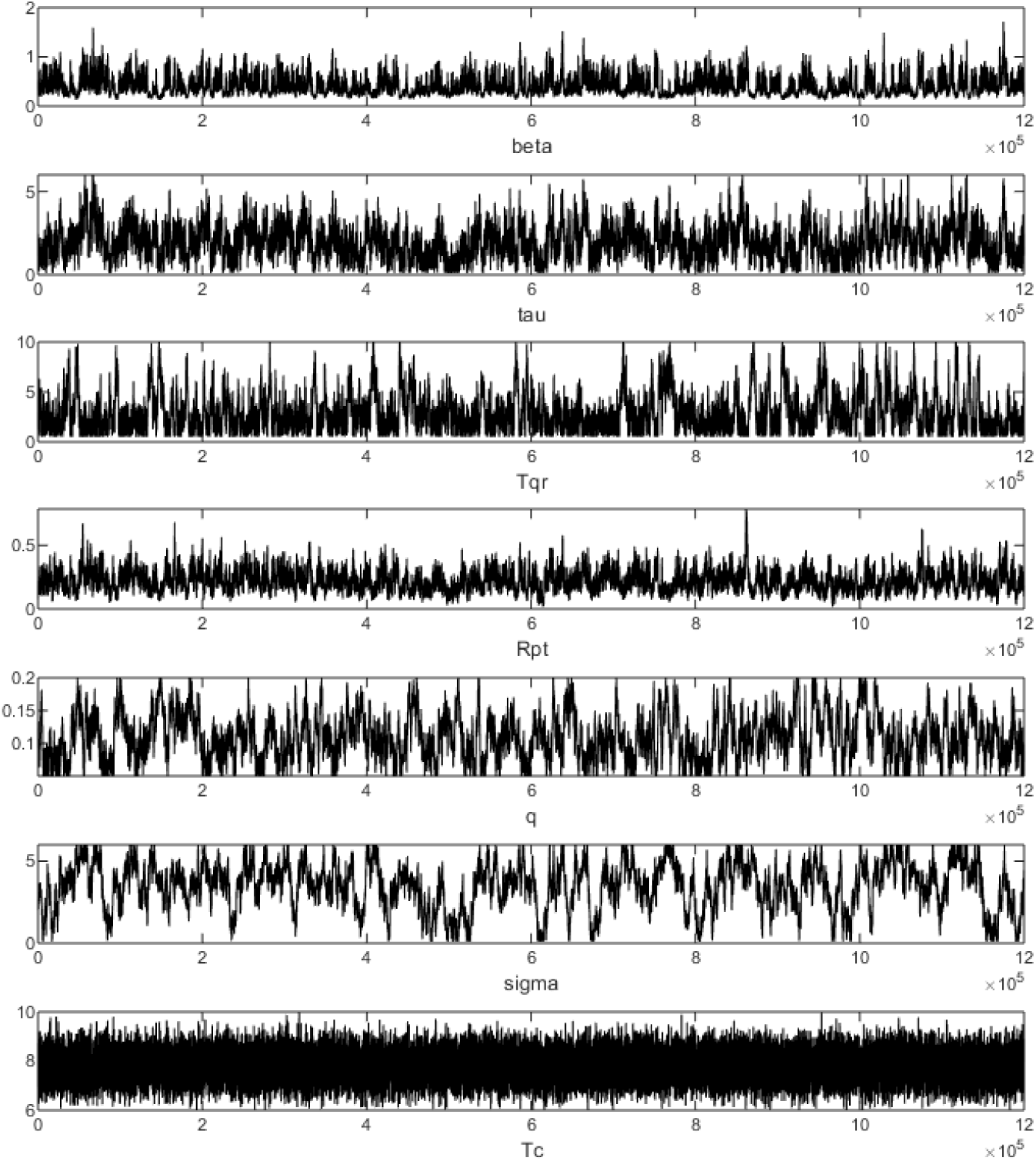
Trajectories of MCMC output.

**Table S1.** Daily number of passengers arriving in Hong Kong.

| Time | Residents(HK) | Residents(Mainland) | Time | Residents(HK) | Residents(Mainland) |
| --- | --- | --- | --- | --- | --- |
| 1/24 | 102,663 | 36,705 | 2/12 | 15,792 | 729 |
| 1/25 | 86,417 | 29,891 | 2/13 | 13,312 | 751 |
| 1/26 | 163,668 | 36,690 | 2/14 | 13,507 | 683 |
| 1/27 | 177,470 | 28,780 | 2/15 | 16,089 | 791 |
| 1/28 | 188,788 | 24,156 | 2/16 | 20,325 | 749 |
| 1/29 | 197,572 | 27,780 | 2/17 | 15,990 | 643 |
| 1/30 | 132,506 | 19,555 | 2/18 | 14,383 | 579 |
| 1/31 | 116,544 | 16,058 | 2/19 | 12,801 | 567 |
| 2/1 | 115,122 | 13,382 | 2/20 | 13,677 | 648 |
| 2/2 | 122,399 | 11,715 | 2/21 | 14,557 | 682 |
| 2/3 | 111,033 | 13,461 | 2/22 | 15,440 | 651 |
| 2/4 | 54,816 | 9,511 | 2/23 | 20,241 | 673 |
| 2/5 | 44,566 | 8,760 | 2/24 | 16,972 | 588 |
| 2/6 | 65,122 | 11,009 | 2/25 | 15,207 | 558 |
| 2/7 | 76,899 | 12,746 | 2/26 | 14,317 | 587 |
| 2/8 | 18,823 | 995 | 2/27 | 13,846 | 702 |
| 2/9 | 24,939 | 956 | 2/28 | 15,248 | 558 |
| 2/10 | 18,408 | 832 | 2/29 | 17,881 | 1,084 |
| 2/11 | 15,953 | 738 |  |  |  |

**Table S2.**
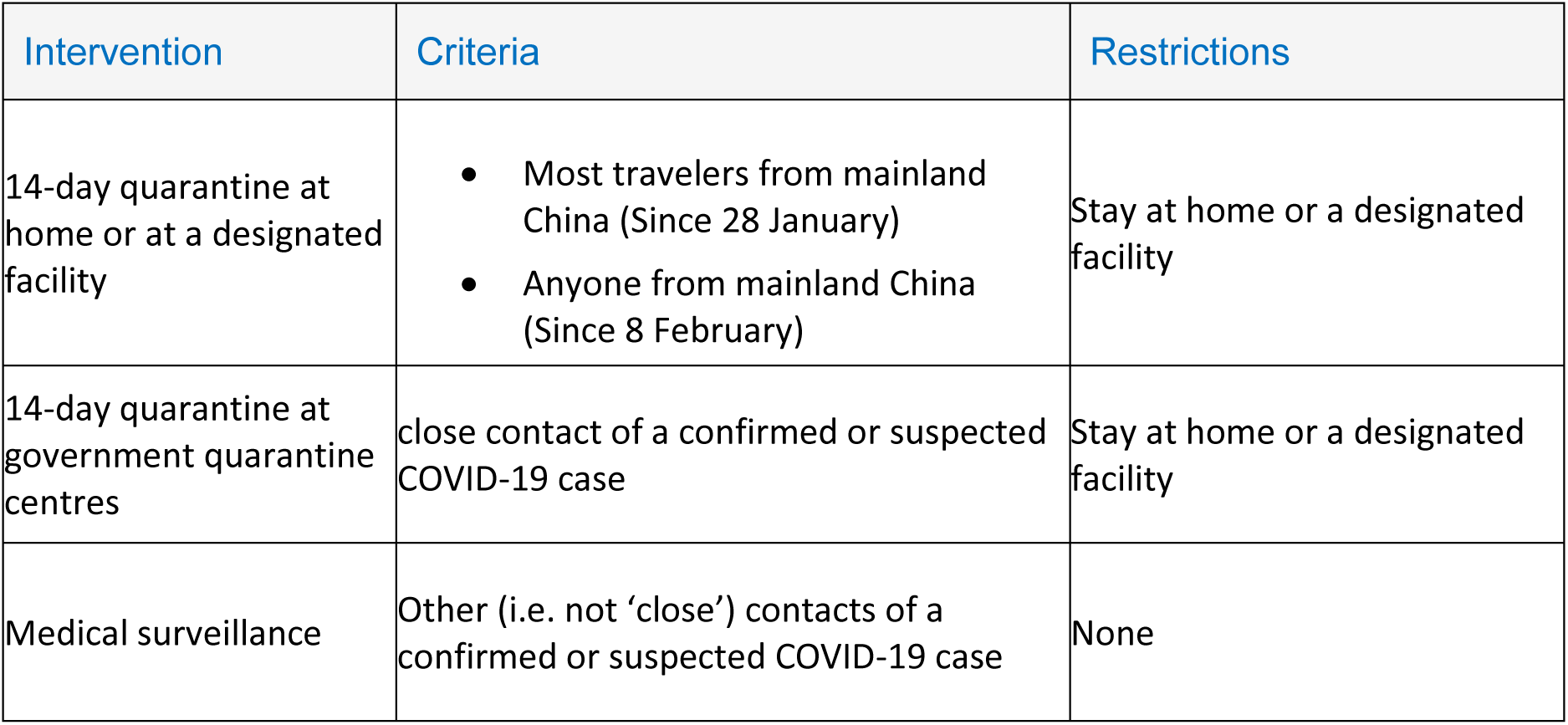
Quarantine-based intervention policies implemented in Hong Kong

